# CoVID-19 in Singapore: Impact of Contact Tracing and Self-awareness on Healthcare Demand

**DOI:** 10.1101/2020.06.04.20122879

**Authors:** Qiuyang Huang, Lin Wang, Yongjian Yang, Liping Huang, Zhanwei Du, Gaoxi Xiao

## Abstract

**Background:** A great concern around the globe now is to mitigate the COVID-19 pandemic via contact tracing. Analyzing the control strategies during the first five months of 2020 in Singapore is important to estimate the effectiveness of contacting tracing measures.

**Methods:** We developed a mathematical model to simulate the COVID-19 epidemic in Singapore, with local cases stratified into 5 categories according to the conditions of contact tracing and self-awareness. Key parameters of each category were estimated from local surveillance data. We also simulated a set of possible scenarios to predict the effects of contact tracing and self-awareness for the following month.

**Findings:** During January 23 - March 16, 2020, the success probabilities of contact tracing and self-awareness were estimated to be 31% (95% CI 28%-33%) and 54% (95% CI 51%-57%), respectively. During March 17 - April 7, 2020, several social distancing measures (e.g., limiting mass gathering) were introduced in Singapore, which, however, were estimated with minor contribution to reduce the non-tracing reproduction number per local case (*R*_*ι*,2_). If contact tracing and self-awareness cannot be further improved, we predict that the COVID-19 epidemic will continue to spread in Singapore if *R*_*ι*,2_ ≥ 1.5.

**Conclusion:** Contact tracing and self-awareness can mitigate the COVID-19 transmission, and can be one of the key strategies to ensure a sustainable reopening after lifting the lockdown.

**Summary:** We evaluate the efficiency of contact tracing and self-awareness in Singapore’s early-stage control of COVID-19. Then use a branching model to simulate and evaluate the possible prospective outcomes of Singapore’s COVID-19 control in different scenarios.

## 1. Introduction

Since the first confirmed case was recorded in December 2019 [1], the disease (COVID-19) has been rapidly spreading across the world [1–3]. With the ongoing global pandemic of COVID-19, the epidemic in Singapore has become increasingly severe. As of 25 May 2020, there were a total of 31,960 confirmed cases and 23 confirmed deaths in Singapore [4]. From the moment the first imported COVID-19 case was confirmed on January 23, Singapore government has been adopting epidemiological investigations and close contact tracing to curb the spread of the coronavirus, aiming to identify individuals who may have been exposed to the confirmed cases, as well as uncovering the links between different cases [4,5].

Contacts tracing is an essential way to control the outbreaks of infectious diseases, such as severe acute respiratory syndrome (SARS) [6], Ebola virus disease [7], Middle East respiratory syndrome (MERS) [8], and many others [9–12]. There is a certain probability that, by effective contact tracing, the infected cases can be isolated before the onset, thereby cutting off the further transmission. In the specific case of COVID-19 where a significant portion of infected have presymptom [13], mild symptoms or no symptoms at all, contact tracing may also help identify such cases after onset and consequently lower their chance of causing further infection spreading.

Individual’s self-awareness of exposure to a confirmed case or cluster is also important for controlling the outbreaks of COVID-19 [14–17]. The uncovered links between different cases and clusters published by the government may help equip the infected individuals with better risk awareness. Consequently, upon observing flu- or pneumonia-like symptoms occurring on themselves, they may tend to more actively seek for medical treatment which helps cut spreading chains earlier. Analysis on surveillance data reveals that earlier confirmation of the first case in a cluster helps reduce the onset-to-isolation delay for the other cases in the same cluster.

Since both close contact tracing and self-awareness of exposure help reduce the delay from onset to isolation [18], and Singapore appeared doing very well in early-stage control by extensive close contact tracing [5], it is worth the efforts assessing the effectiveness of these measures and to what extents they may help control the outbreak of COVID-19. In this paper, by exploring the public data made available in government press releases, we estimate the serial interval distribution and the delay distribution between different states of a case, e.g., from being infected to onset, from onset to isolation and from isolation to confirmation etc. By adopting the estimated distributions and parameters, a stochastic transmission model [19–21] was built to simulate the outbreak of COVID-19 in Singapore, which allowed us to evaluate the effects of contact tracing and self-awareness in early-stage disease control. Varying the efficacy of close contact tracing, self-awareness and the reproduction number in the model, we could also simulate possible future developments of COVID-19 control in Singapore in a few different scenarios.

## 2. Method

### 2.1. Date description

Detailed information of confirmed COVID-19 cases is collected by the Ministry of Health (MOH) in Singapore [2]. Since the confirmation of the first COVID-19 case on 23 January 2020, MOH has publicly reported the details of each confirmed case, including key epidemiologic timelines (e.g., dates of symptom onset, confirmation, and isolation). Imported cases were also reported with their dates of arrival in Singapore. The contacts of each confirmed case were identified by epidemiological investigations and contact tracing. As of 16 April 2020, MOH has reported 247 confirmed cases that formed into 65 clusters. Since March 17, 2020, however, the reported information of each case no longer included such details, probably due to the significantly increased number of cases. Since 2 April 2020, the number of confirmed cases has been substantially increasing among migrant workers living in crowded dormitories. Considering the potential differences in contact and travel behaviors between the migrant workers and local population in Singapore, we excluded those cases among migrant workers from the analysis in this article.

### 2.2. Simulation model

Basic reproduction number *R*_0_ denotes the expected number of secondary infections caused by an infector in a fully susceptible population, which is suitable to the early-stage of COVID-19 epidemic in Singapore (January 23 – March 16, 2020). During this stage, all 54 infectors detected by MoH were imported from other countries, and contact tracing during this stage mainly aimed to quarantine infectees caused by imported cases. We defined the non-tracing reproduction number as the expected number of secondary infections caused by an infector given the use of certain non-pharmaceutical control measures (e.g., social distancing) other than contact tracing and self-awareness, and the effective reproduction number as the reproduction number when contact tracing and self-awareness are combined with other non-pharmaceutical control measures.

We used a stochastic branching process model to simulate the COVID-19 epidemic in Singapore (Figure 2). Each potential infector may infect a number of secondary cases, which follows a negative binomial distribution with a mean equaling the non-tracing reproduction number [22,23] (Figure 3A). The symptom onset time of each secondary case is determined by the serial interval distribution. And the infectee’s generation time is determined by the time lag between the infectee’ symptom-onset time and the incubation period [24,25] (Figure 3B). We estimated the serial interval distribution by using 72 pairs linked cases, following a skewed normal distribution (Figure 3C). We also estimated the time delays between different states (onset to isolation, isolation to confirmation, etc.) by using the surveillance data (Figures 3D-F, details in next subsection), such estimations are of critical importance to simulate the COVID-19 epidemic. Table 1 summarizes all parameters. Using the information about imported cases, we nowcast the cumulative number of confirmed COVID-19 cases by simulating the branching process seeded by each imported case. To account for the multiple non-pharmaceutical control measures implemented in Singapore during different stages of the COVID-19 epidemics, we simulated the branching processes seeded by imported or local cases using different types of reproduction numbers (Table 1).

**Figure 1:**
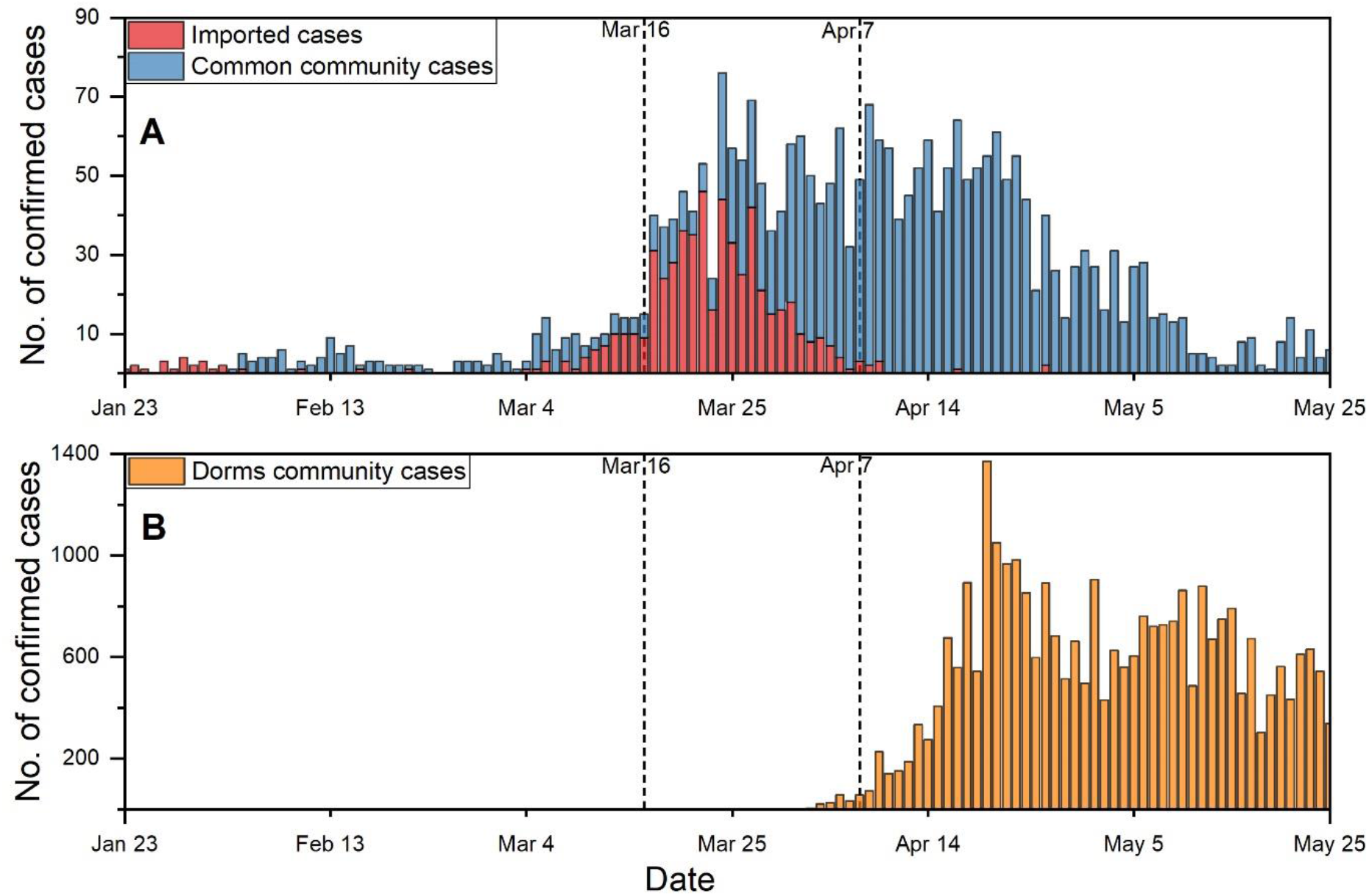
Daily number of confirmed COVID-19 cases by confirmation dates. We considered the COVID-19 epidemic in Singapore as three stages, divided by dashed lines (March 16 and April 7), because different control measures were taken at different stages. (A) The number of new confirmed COVID-19 cases each day, who were imported from other countries (colored in red) or locally infected in Singapore (colored in blue). (B) The number of new confirmed COVID-19 cases each day among migrant workers.

**Figure 2:**
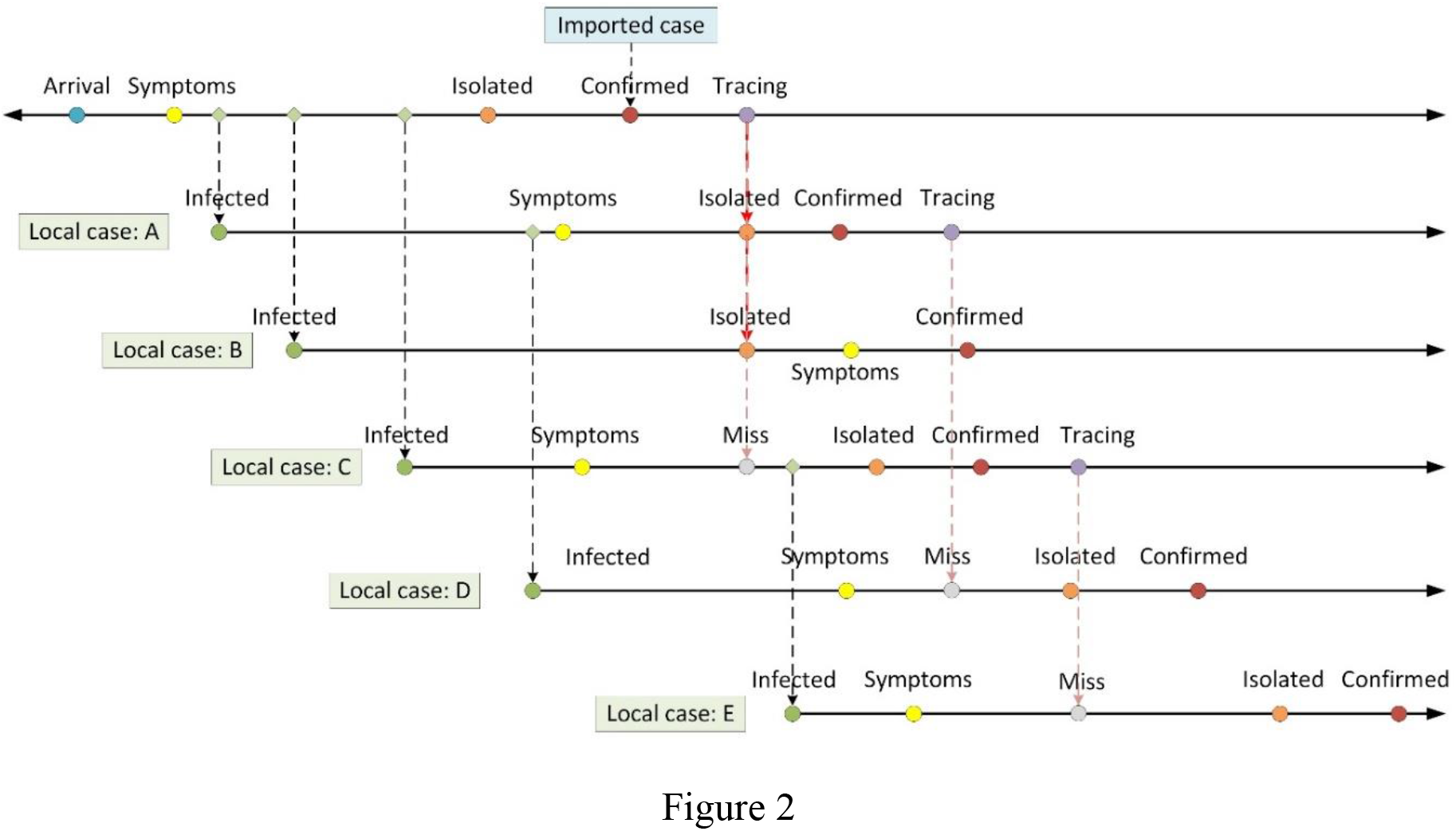
Schematic of a cluster of 6 COVID-19 cases simulated with branching process model. This cluster is seeded by a single imported case, which generates three local cases (local cases A, B and C), then local transmission starts (case A generated case D, and case C generated case E). Each line indicates a case, with each colored circle indicates the change in infection status. Line segment between olive (infection time) and yellow (symptom onset time) circles indicates the incubation period, line segment between yellow and orange (isolation time) circles indicates the time delay from symptom onset to isolation, line segment between orange and red (confirmation time) circles indicates a short delay from isolation to confirmation (which is set to zero if the case is confirmed before isolation), line segment between red and violet (time of tracing) circles indicates the tracing delay. Each case is detected as a close contact of a confirmed case with probability *tp*, which leads to the isolation of the detected case (with a tracing delay after his/her infector was confirmed.

**Figure 3:**
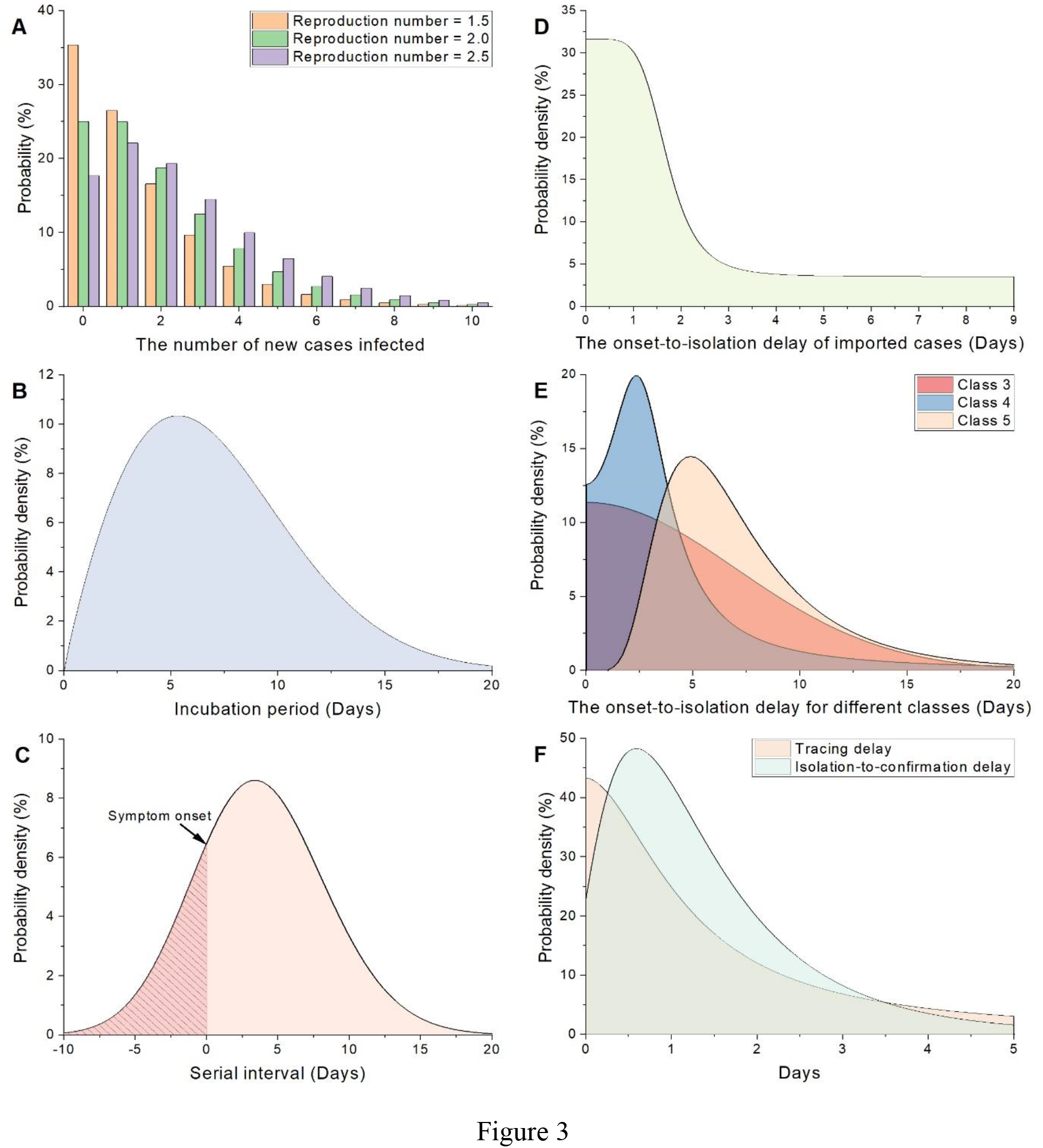
Probability distributions used in simulations. (A) The distribution of the number of potential secondary cases infected by each individual, drawn from a negative binomial distribution, of which the mean equals the non-tracing reproduction number. We illustrate 3 different cases with different reproduction number values as examples. (B) The incubation period distribution in Singapore estimated by Lauren C et al. [24] (C) Serial interval distribution fitted by skewed normal distribution. Here we set 0 as the symptom onset time. (D) The delay distribution from onset to isolation of imported cases. Since there is only information of the confirmed date of each case after March 16, we have to estimate the symptom onset date for each imported case. (E) The delay distribution from onset to isolation of Classes 3–5. (F) The distribution of tracing delay and isolation to confirmed delay. The distributions in (C), (D), (E) and (F) were all fitted by using surveillance data. Details of the estimations of the distributions and parameters were given in the Supplementary Materials.

**Table 1:** Model Parameters.

| Parameter | Mean (95% CI) | Distribution | Source |
| --- | --- | --- | --- |
| The number of new cases infected by each individual | Reproduction number | Negative binomial distribution | Joel Hellewell et al [18] |
| Incubation period | 7.1 days (6.13, 8.25) | Weibull distribution | Lauren C et al [20] |
| Serial interval distribution | 3.23 days (2.57, 4.16) | Skewed normal distribution | Estimated |
| Onset-to-isolation delay for imported cases | 1.87 days (1.36, 2.39) | Logistic distribution | Estimated |
| Onset-to-isolation delay for Class 3 | 5.77 days (4.10, 7.44) | Skewed normal distribution | Estimated |
| Onset-to-isolation delay for Class 4 | 4.17 days (2.43, 5.91) | Folded Cauchy distribution | Estimated |
| Onset-to-isolation delay for Class 5 | 7.84 days (6.23, 9.45) | Exponentiated Weibull distribution | Estimated |
| Tracing delay for Classes 1 and 2 | 1.42 days (0.88, 1.95) | Logistic distribution | Estimated |
| Isolation-to-confirmation delay | 1.44 days (1.25, 1.63) | Lognormal distribution | Estimated |
| Contacts tracing probability $tp$ from Jan 23 to Mar 16 | 0.31 (0.28, 0.33) | -- | Estimated |
| Self-awareness probability $ap$ from Jan 23 to Mar 16 | 0.54 (0.51, 0.57) | -- | Estimated |
| Basic reproduction number of all cases $R_0$ | 2.1 | -- | Estimated |
| Non-tracing reproduction number of local cases from Mar 16 to Apr 7: $R_{l,2}$ | 1.9 | -- | Estimated |
| Non-tracing reproduction number of imported cases from Mar 16 to Mar 25: $R_{i,2}$ | 1.5 | -- | Assumed |
| Non-tracing reproduction number of imported cases after Mar 25: $R_{i,3}$ | 0 | -- | Assumed |
| Non-tracing reproduction number of local cases after Apr 7: $R_{l,3}$ | 1.5-2.1 | -- | Tested |

\*We used scipy (a package of python) to fit the distributions, the parameters were given in Supplementary Table 2 and 4.

We use the following two parameters to describe the effects of contact tracing and self-awareness: (1) contact tracing probability (*tp*); and (2) self-awareness probability (*ap*). We assume that each secondary case can be successfully traced as a close contact with a probability *tp*. The contact tracing started with a short delay (tracing delay) after a case was confirmed; once a secondary case was traced, he will be isolated immediately (such as the cases A and B in Figure 2). Here we assumed that isolations, once imposed, were 100% effective in preventing further transmission. For those cases that were missed by contact tracing (with a probability 1 − *tp*), they may have self-awareness of exposure (at a probability *ap*). Both contact tracing and self-awareness may reduce time delay from symptom onset to isolation (i.e., isolation delay).

We categorized local cases of each transmission cluster into the following five classes and estimated their respective isolation delay distribution from surveillance data (details in next subsection):

- Class 1: Detected close contacts (of any confirmed case) who were isolated before developing symptoms (Figure 2, local case B).
- Class 2: Detected close contacts (of any confirmed case) who were traced and isolated after developing symptoms (Figure 2, local case A).
- Class 3: Not detected as close contacts of any confirmed case and not isolated before developing symptoms. Such individuals may recognize their potential exposure by self-awareness. Their symptom onsets occurred earlier than the confirmation of the first case in that cluster, so their self-awareness occurred after developing symptoms.
- Class 4: Similar to Class 3, except that their symptom onsets occurred later than the confirmation of the first case in that cluster.
- Class 5: Not detected as close contacts of any confirmed case and no self-awareness after developing symptoms (Figure 2, local case E).

Cases of Class 1 and Class 5 have the lowest and highest chances to infect others. Cases of Class 2, 3, 4 have a reduced probability of infecting others due to contact tracing and self-awareness.

## 3. Results

### 3.1. Parameters estimation

The MoH reported 247 confirmed COVID-19 cases in Singapore before March 16, 2020, including 89 cases imported from other countries, 15 local cases of Class 1, 52 local cases of Class 2, 25 local cases of class 3, 29 local cases of class 4, and 37 local cases of Class 5. From this data, we reconstructed 72 transmission pairs, in which the symptom-onset times were available for both the infector and infectee. This allows us to compute their empirical serial intervals. For each of 247 confirmed cases, we also computed their time delays between different infection status, which were used to fit the probability distributions used in our simulation model. Specifically, the cases of Classes 1 and 2 were correctly identified as close contacts, which were isolated with a short delay (tracing delay) after their infectors were diagnosed. We counted the tracing delay of these 67 cases. We also counted the delay from onset to isolation for each case in Classes 3 to 5 and the imported cases, as well as the isolation to confirmation delay of all the 247 cases.

We estimated the distributions for 7 types of time delays, including the serial interval, tracing delay, onset to isolation delay for 4 different classes, and isolation to confirmation delay for all cases via Maximum Likelihood Estimation. We used the Mean Square Error (MSE) to select candidate models that best fit the distributions of time delays. The best-fit distribution of each time delay with estimated parameters were summarized in Table 1, with probability distributions shown in Figures 2C-F. Details on parameter estimations are given in Supplementary Materials.

Based on MoH case series data and our estimated parameters, we estimated that the mean success probability of contact tracing *tp* is 0.31 with 95% confidence interval (CI) as (0.28, 0.33), and the mean success probability of self-awareness ap is 0.54 with 95% CI as (0.51, 0.57) (Figure 4). Finally, by varying *R*_0_ from 1.5 to 2.5, which spans most of the ranges reported by current estimates [26–29], we estimated the basic reproduction number for both imported cases and local cases from January 23 to March 16 as *R*_0_ = 2.1 (see Supplementary Figure 4).

**Figure 4:**
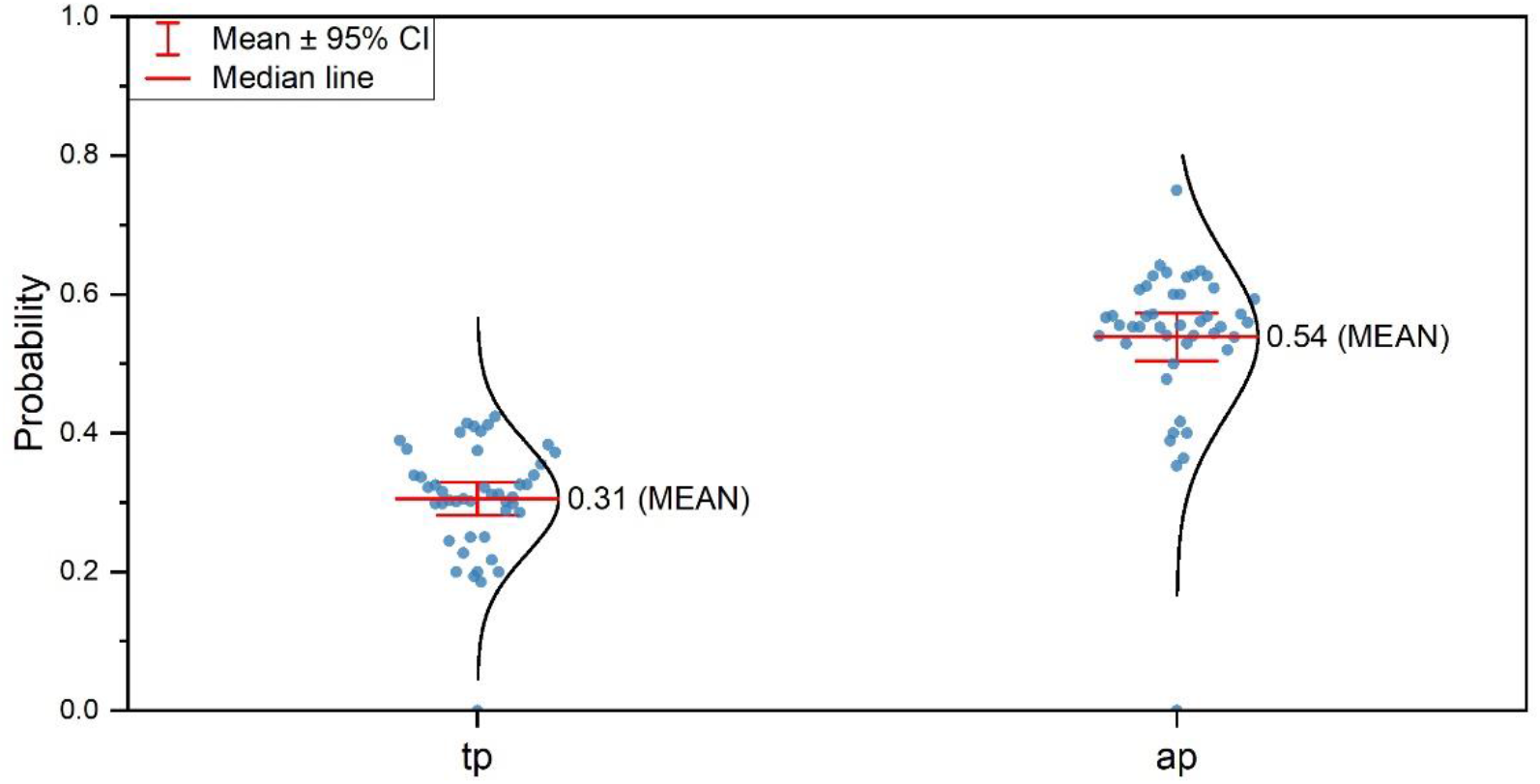
The probability of successful contact tracing and self-awareness. The blue dots represent the probability in each day from January 23 to March 16, the black lines represent the norm distribution, and the red lines represent the mean value and the 95% CI.

There were several limitations in our data processing. We used the data published by MOH in press release to infer whether a case was traced and isolated; a few cases may be misclassified. Due to the lack of detailed information since March 17, 2020, only 247 cases can be analyzed. For all cases not explicitly stated as being isolated upon onset, we assumed that they were not being isolated at that moment. This might lead to an underestimation of Class 2 and an overestimation of Classes 3 and 4. Lastly, effects of asymptomatic cases are not taken into account in this study, mainly due to the lack of data. However, there was evidence indicating that the effects of asymptomatic cases may not be significant in Singapore [30].

### 3.2. Simulation scenarios

We considered the COVID-19 epidemic in Singapore as three stages. Stage 1 was between 23 January 2020 and 16 March 2020, during which the MoH provided detailed reports for each of the 247 confirmed COVID-19 cases (including 54 imported cases). We used these reports to estimate the basic reproduction number (*R*_0_) and the success probabilities of contact tracing (*tp*) and self-awareness (*ap*).

Stage 2 was between 17 March 2020 and 7 April 2020, during which 469 COVID-19 cases were imported into Singapore (more than sevenfold higher than that of stage 1). As such, tighter restrictions (e.g., strict restriction of non-essential travel to countries affected by COVID-19, 14-day stay-home quarantine for returning residents) started to be implemented upon travelers. For imported cases, we assumed that (1) the non-tracing reproduction number (*R*_𝑖,2_) was 1.5 [31] during March 17 – 24, 2020, and (2) was reduced to *R*_𝑖,3_ = 0 from 25 March 2020 because all travelers from other countries were required to do 14-day quarantine after arrival. For local cases, additional social distancing measures, e.g., limiting mass gathering, increasing space among individuals in public places, etc., (announced on March 13 and effected on March 16) were implemented. Because of the lack of granular data on confirmed cases for estimating parameters during stage 2, we used the same success probabilities of contact tracing (*tp*) and self-awareness (*ap*) as those of stage 1 to simulate the transmission of COVID-19, with the aim of evaluating the reduction in non-tracing reproduction number of local cases (*R*_ι,2_) due to the social distancing measures implemented during stage 2.

Stage 3 started from 7 April 2020, during which the government implemented a stringent set of preventive measures termed ‘Circuit Breaker Measures (CBM)’, e.g., stay at home as much as possible, controlled access at areas susceptible to crowding, closure of more work premises and tighter workplace measures, etc. At the time of writing, Singapore is still implementing the CBM. Although granular data on confirmed cases is still not available for this stage, we can estimate the control effect based on our results on the stages 1 and 2. We tested several possible scenarios on the success probabilities of contact tracing and self-awareness for stage 3, and estimated the extent of social contacts that need to be reduced to achieve effective disease control.

### 3.3. Effect of contact tracing and self-awareness in mitigating the spread of COVID-19

By modelling the effect of contact tracing and self-awareness with parameters estimated for the data of stage 1, we found that our simulated epi-curve of cumulative incident cases is consistent with that reported by MoH (Figure 5A). To evaluate the effect of contact tracing and self-awareness during stage 1, we also performed simulations by excluding contact tracing or self-awareness or excluding both of them. Comparison results presented in Figure 5A demonstrated the effectiveness of the contact tracing and self-awareness in preventing an outbreak in the first stage. We can observe that, without contact tracing or self-awareness or without both of them in simulation, the number of total confirmed cases at the end of stage 1 would be 317, 354 and 431 respectively, which is significantly higher than ground truth (243 confirmed cases).

**Figure 5:**
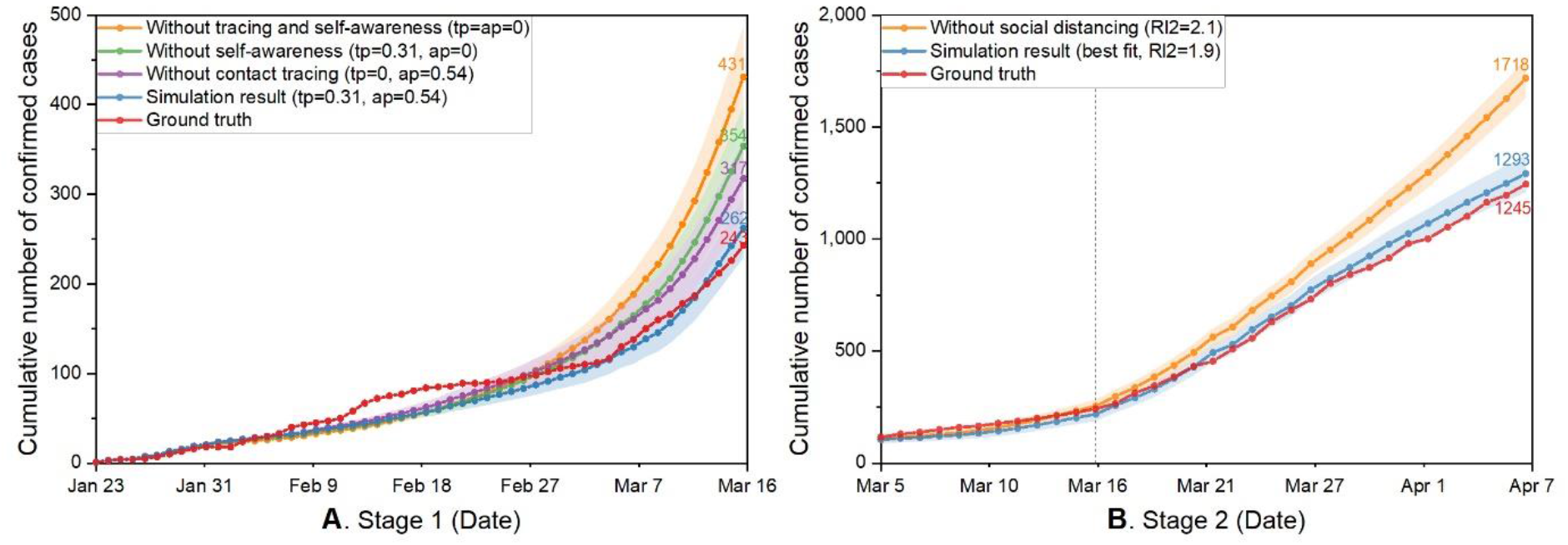
Simulating the effect of contact tracing and self-awareness for stages 1 (January 23 - March 16) and 2 (March 17 - April 7) (A) Simulations with or without contact tracing and self-awareness for the stage 1. Excluding contact tracing or excluding self-awareness in simulations will increase the total number of confirmed cases at the end of stage 1 by about 30% and 46%, respectively. Excluding contact tracing and self-awareness together, there would be 431 confirmed cases by the end of stage 1, which is more than 1.7-fold higher than that reported by MoH. (B) Simulations with or without social distancing measures for stage 2. If social distancing measures had not been adopted, $$Rl2$$ had remained as 2.1, as can be observed that, there would have been a more drastic increase leading to an infection size about 40% larger.

With the entry of a large number of imported cases in stage 2, the confirmed cases increased substantially (Figure 5B). The social distancing measures adopted by the government helped reduce the non-tracing reproduction number in this stage. Consequently, the cumulative curve approximately still had a linear growth rate. By assuming that the contact tracing and self-awareness remained as effective as before, we found that a non-tracing reproduction number of *R*_*ι*,2_ = 1.9 leads to the minimal errors. Considering that self-awareness probably became less effective in stage 2 due to an increasing number of cases and an overwhelming flow of information, the effects of social distancing may have been actually more effective than lowering *R*_*ι*,2_ to 1.9. A linear increase at a fast rate however would almost certainly degrade the efficiency of contact tracing and self-awareness [5]. The conclusion hence has to be that further control schemes (e.g., CBM) need to be introduced.

As stage 3 is currently ongoing, as aforementioned, we only tested on a few different cases with representative parameter values. Different cases were tested where (1) both contacting tracing and self-awareness remain as effective as before; (2) contact tracing remains as effective as before yet self-awareness is degraded; and (3) both are degraded respectively. We see in Figure 6 that *R*_*ι*,3_ has to be lower than 1.5, 1.4 and 1.3 respectively in order to put the disease spreading under control. In real life, it may be very difficult to keep the non-tracing reproduction number of a strongly infectious disease as COVID-19 to be as low as 1.3 without a major lockdown. Hence the observations may indicate that contact tracing and self-awareness have to be reasonably effective in order to contain disease spreading while avoiding a major lockdown.

**Figure 6:**
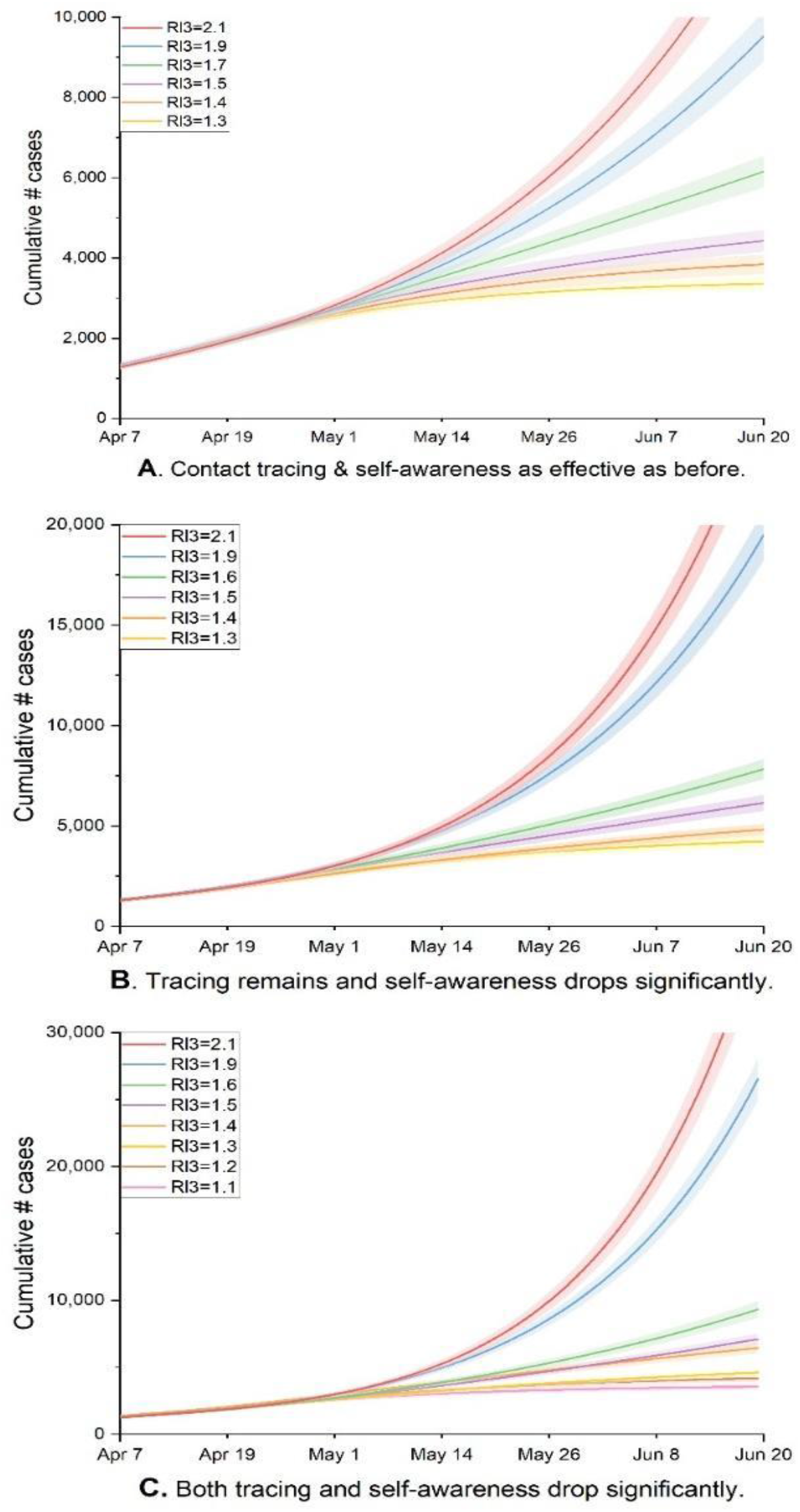
Simulation results under different scenarios in stage 3. (A) For scenarios where both contact tracing and self-awareness remain as efficient as before (*tp* = 0.31, *ap* = 0.54). To put disease spreading under control, Rl3 has to be lower than 1.5. (B) For scenarios where self-awareness degrades yet contact tracing remains as efficient as before (*tp* = 0.31, *ap* = 0.1). with *R*_ι,3_ ranging from 1.3 to 2.1, the infection size would be increased by 1.2 to 2.1 times by June 20 compared to the corresponding ones in Figure 6A. To keep cumulative cases increase at a linear rate or lower, *R*_ι,3_ needs to be lower than 1.4. (C) For scenarios where performances of both contact tracing and self-awareness degrade significantly (*tp* = *ap* = 0.1). The infection size will be increased by about 3 times compared to those in Figure 6A. To have a linear or lower increasing rate in cumulative cases, *R*_ι,3_ needs to be lower than 1.3.

## 4. Discussion

By analyzing the 247 cases in Singapore with relatively detailed data, we estimated the average efficiency of contact tracing and self-awareness in early stage. By dividing the local cases into five different classes. We found that in addition to cutting off infection spreading from those cases in Class 1 altogether, close contact tracing and self-awareness helped reduce the delay from onset to isolation by an average of 2–4 days for those cases belonging to Classes 2–4, which helped reduce the average number of secondary cases.

Based on our model, we estimated the basic reproduction number in Singapore to be likely between 2 and 2.5 unless there is a large portion of asymptomatic cases. This is evidenced by the reasonably good match between simulation results and ground truth. By introducing social distancing in stage 2, the non-tracing reproduction number (*R*_*ι*,2_) may have been reduced to be about 1.9 or slightly lower.

Singapore’s experiences show that close contact tracing and self-awareness combined together may achieve effective control of the epidemic spreading when the number of infected cases is low. A booming number of imported cases, together with a degraded performance of contact tracing and self-awareness such a booming number may easily lead to, may quickly drive disease spreading from being under control to a dangerous outbreak. It is shown that, in order to stop the disease spreading in stage 3, CBM and other control schemes need to be imposed to push down the non-tracing reproduction number (*R*_ι,3_) to be lower than 1.5 if contact tracing and self-awareness remain as effectively as before, or even lower with degraded contract tracing and self-awareness.

The above discussions indicate that contact tracing and self-awareness must be enhanced as much as possible when lockdown comes to an end. Some plausible efforts have been made in Singapore to improve the accuracy of contact tracing, e.g., the Bluetooth-based “TraceTogether” app [32–34] for close contact tracing. Other tracing schemes, e.g., IC scanning at supermarket entries are being imposed. More of such schemes are needed, while a tricky balance between effective tracing and minimum intrusion into privacy has to be kept. Studies on such topics are urgently needed

As aforementioned, the current simulation results are based on the assumption that the asymptomatic cases do not play a significant role in the disease spreading and control. When more knowledge of the scale and infectivity of asymptomatic cases is available, the proposed model may be adjusted to provide more accurate estimation and prediction results. Such adjustments could be done in a relatively straightforward manner.

## Notes

### Author contributions

Gaoxi Xiao conceived the study. Qiuyang Huang and Gaoxi Xiao designed the model and wrote the article. Liping Huang and Yongjian Yang worked on statistical aspects of the study. All authors reviewed and approved the final version of the manuscript.

## Data Availability

Consisting of data provided by the authors to benefit the reader, the posted materials are not copyedited and are the sole responsibility of the authors, so questions or comments should be addressed to the corresponding author.

## Acknowledgements and financial support

The authors thank Prof. Benjamin Cowling for helpful discussions. The project is partially supported by the Open Fund of Key Laboratory of Urban Land Resources Monitoring and Simulation, Ministry of Land and Resources, China (No. KF-2019–04–034), and National Research Foundation, Singapore, under contract NRF2017VSG-AT3DCM001–045. QH gratefully acknowledges financial support from China Scholarship Council.

